# Factors Associated with Delayed Hospital Arrival after Stroke Onset: An Observational Study in Thanh Hoa Province, Vietnam

**DOI:** 10.1101/2024.01.29.24301925

**Authors:** Hoa Thi Truong, Sam Hoanh Nguyen, Cuong Van Le, Shinichi Tokuno, Aya Kuchiba, Shinji Nakahara

## Abstract

**Background:** Delayed hospital arrival lowers the proportion of patients with stroke receiving recanalization therapy and results in poor outcomes. This study investigated the factors associated with pre-hospital delays in hospital arrival after stroke onset in the Thanh Hoa Province, Vietnam.

**Methods:** Clinical data were collected from stroke patients within 7 days of symptom onset who were prospectively registered in this study. Patients and/or their relatives were interviewed using a structured questionnaire about patient social demographics, address, post-stroke support actions, and stroke awareness. Pre-hospital delay in hospital arrival was dichotomized into ≤ 4.5 hours and > 4.5 hours, and multivariable logistic regression analysis was used to investigate factors associated with the delay.

**Result:** Of the 328 participants analyzed, 181 (55.4%) arrived at the hospital 4.5 hours after the symptom onset. The patients’ and relatives’ awareness of stroke was poor. Pre-hospital delays were longer for patients living > 10 km away from a healthcare facility and those with secondary or lower education levels, with odds ratios of 2.07 and 1.98, respectively. Seeking care at a district or private hospital as the first point of healthcare or non-use of emergency medical services did not show significant associations.

**Discussion:** The study revealed that most patients with stroke did not arrive at the hospital in time for recanalization therapy. Moreover, the low stroke awareness among patients and their relatives is concerning. Further research is needed to investigate the reasons for pre-hospital delays and develop targeted interventions to improve stroke awareness and reduce these delays.

## INTRODUCTION

Stroke is the second major cause of global death and the third most common cause of disability-adjusted life years, with a rapidly rising incidence in low- and middle-income countries (LMICs). (1) Stroke is also a noteworthy health issue in Vietnam, with approximately 200,000 cases reported annually, 50% of which are not appropriately treated. (2, 3) Recanalization therapy (including thrombolysis and endovascular thrombectomy) can improve patients’ prognosis; however, is highly time-dependent. (4–6) Despite rigorous media campaigns to raise people’s awareness of the urgent medical seeking required when a stroke is suspected, delayed hospital arrival is still common among patients with stroke. (7–9) The median time from stroke onset to hospital presentation ranged between 4 h and 24 h in previous studies from Australia and Sweden. (9–11)

Several studies have investigated this matter to pinpoint the factors associated with the delays. They have highlighted that such factors encompass inefficient handling of hospitalization paperwork, referrals, and the time taken to diagnose the patient and commence treatment. (11–13) Additionally, common factors associated with pre-hospital delay include the lack of awareness of stroke symptoms among patients and their relatives, low education level, long distance from a hospital, and face-to-face visits to their family doctors. (14–18)

The majority of these studies have been conducted in high-income countries (HICs) with established emergency medical services (EMS) systems. At the same time, there remains a lack of research in LMICs with underdeveloped EMS infrastructure. Furthermore, variations in geography, ethnicity, culture, demographics, and healthcare systems across countries pose challenges to the generalizability of findings from prior studies.

The objective of this study was to characterize pre-hospital delays in hospital arrival as well as delays in conducting hospital procedures. Additionally, the study aimed to examine factors associated with the pre-hospital delays, focusing on using EMS to transport patients to the hospital and the option of going directly to a specialized hospital.

## METHODS

### Study settings

Thanh Hoa General Hospital is a tertiary care public hospital with 1200 hospital beds, located in the Thanh Hoa Province, spanning an area of 11,168.3 km^2^, and comprising a population of approximately 3,640,000. Its neurology department is the only facility in the province capable of performing thrombolysis and mechanical thrombectomy in patients with stroke, who are usually admitted to the emergency department and transferred to the neurology department for treatment after diagnosis. A few patients with stroke are, however, directly transported to the neurology department.

Under the Ministry of Health, Vietnam’s national public EMS service can be accessed by dialing the nationally uniform phone number 1-1-5. Each province must have an ambulance dispatch center, known as a 1-1-5 center. Thanh Hoa Province’s EMS center is located at the Thanh Hoa General Hospital, dispatching emergency response ambulances. However, district hospital ambulances are primarily used for patient referrals between hospitals, and the province’s current EMS coverage is inefficient, leading to patient reluctance to utilize the service.

### Study design

This observational study was conducted at the Neurology and Stroke Department of Thanh Hoa General Hospital from April 1, 2022, to July 10, 2022. Clinical data were collected from stroke patients within 7 days of symptom onset who were prospectively registered in this study. Among the patients, the time intervals from symptom onset to emergency department arrival and to the neurology department for definitive treatment were investigated, and factors associated with pre-hospital delay were determined. Written informed consent was obtained from the patients or their relatives. The study was approved by the appropriate ethics committees in both Vietnam and Japan.

### Participants

Patients were included if they met all the criteria, as shown in **Table 1**. Inclusion criteria were patients with (1) acute focal neurological symptoms that occurred outside the hospital, including wake-up patients with stroke, (2) lesions corresponding to CT or MRI/A, and (3) those seeking medical help at the emergency department of Thanh Hoa General Hospital (arrival time could not be determined in other departments) within 7 days from symptom onset. Patients referred from another hospital after receiving any definitive treatment were excluded because such patients were referred with worsening conditions after treatment.

**Table 1.**
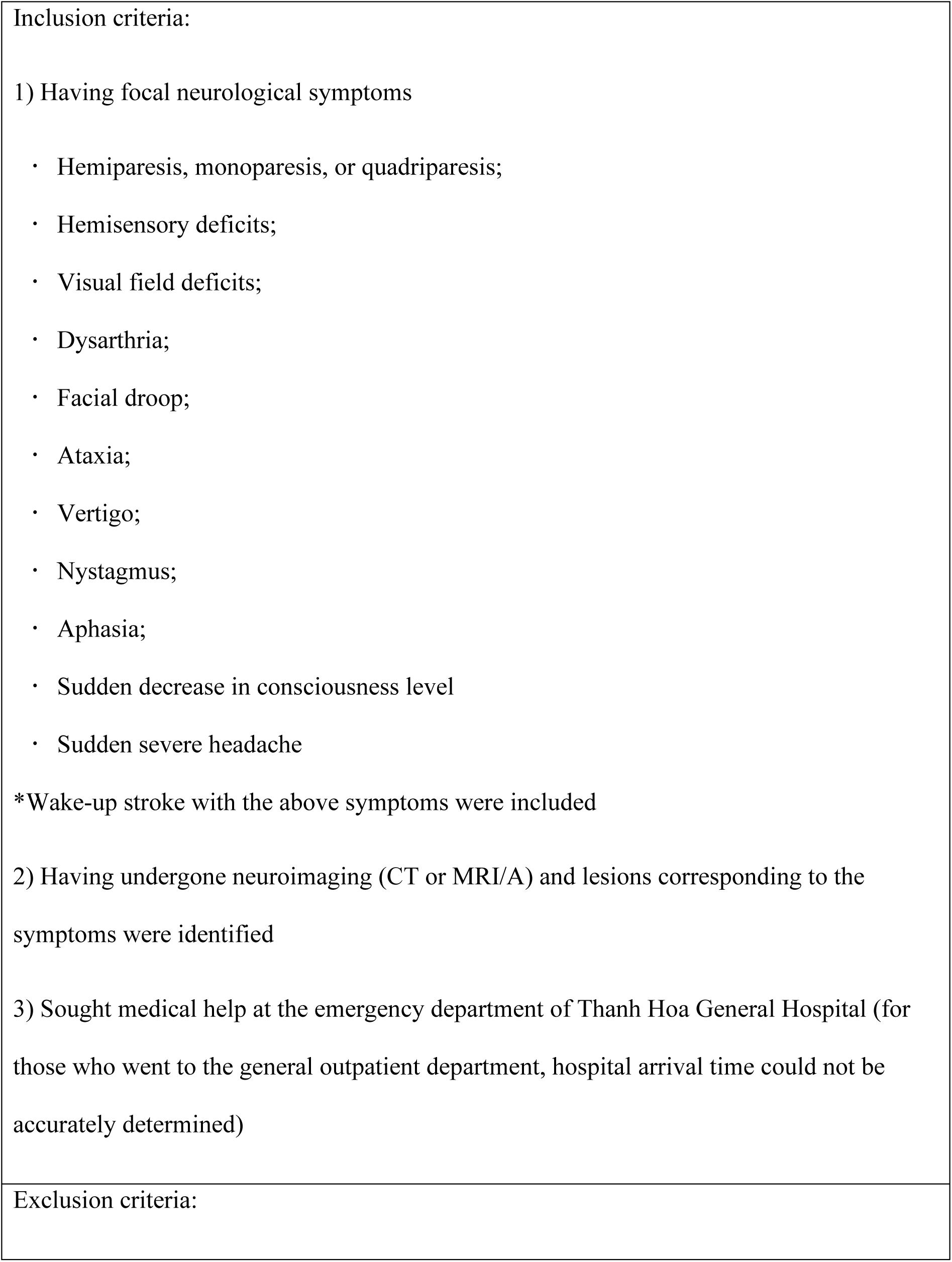
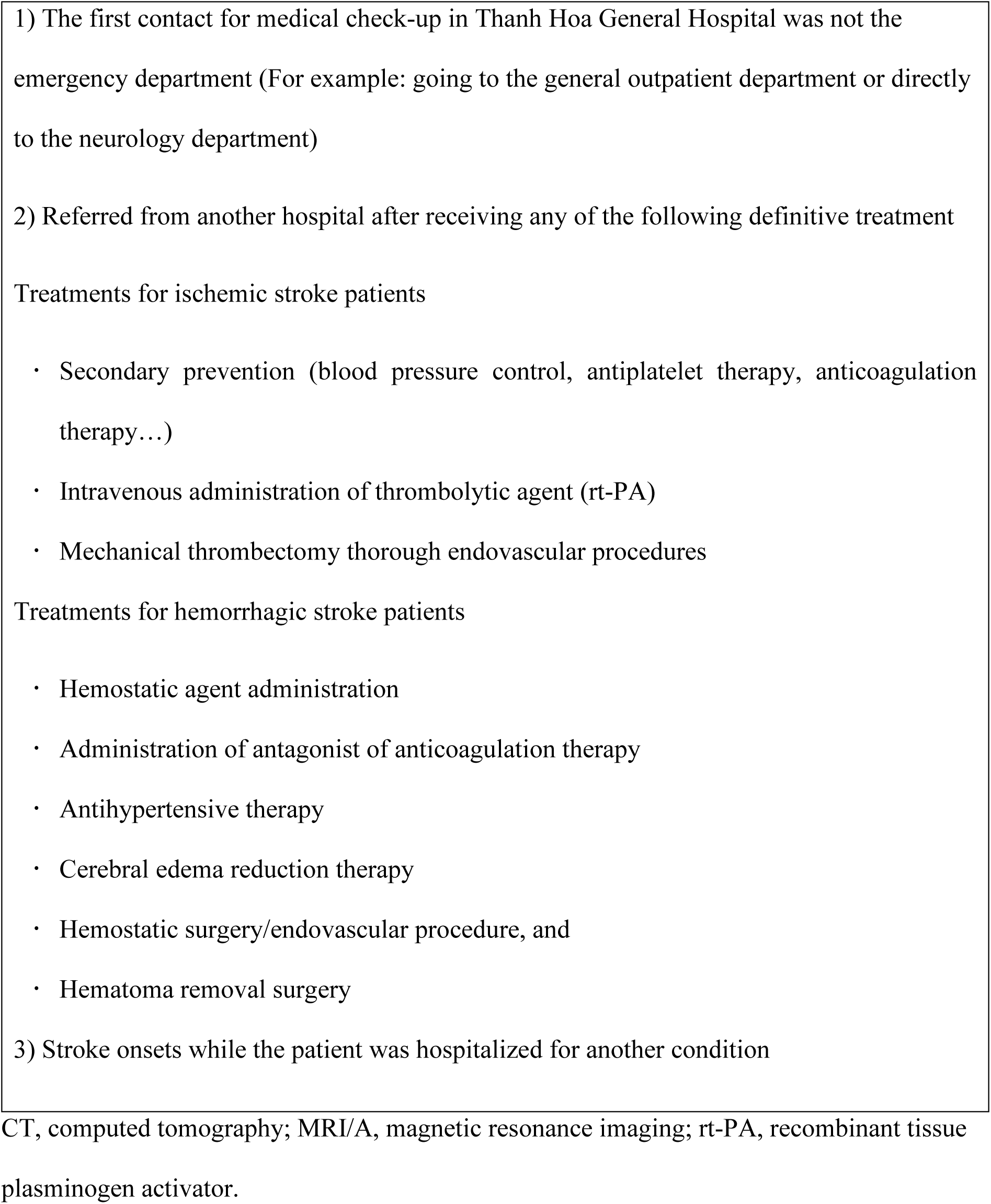
Inclusion and exclusion criteria.

Out of the 346 eligible patients at the time of admission, 15 were excluded from the study as they were unavailable for interview due to referral, discharge, or death within 1–2 days after admission. In addition, a mismatch between the medical records and interview data files occurred in three patients, who were also excluded (**Figure 1**). Thus, 328 eligible patients were included in the study.

**Figure 1.**
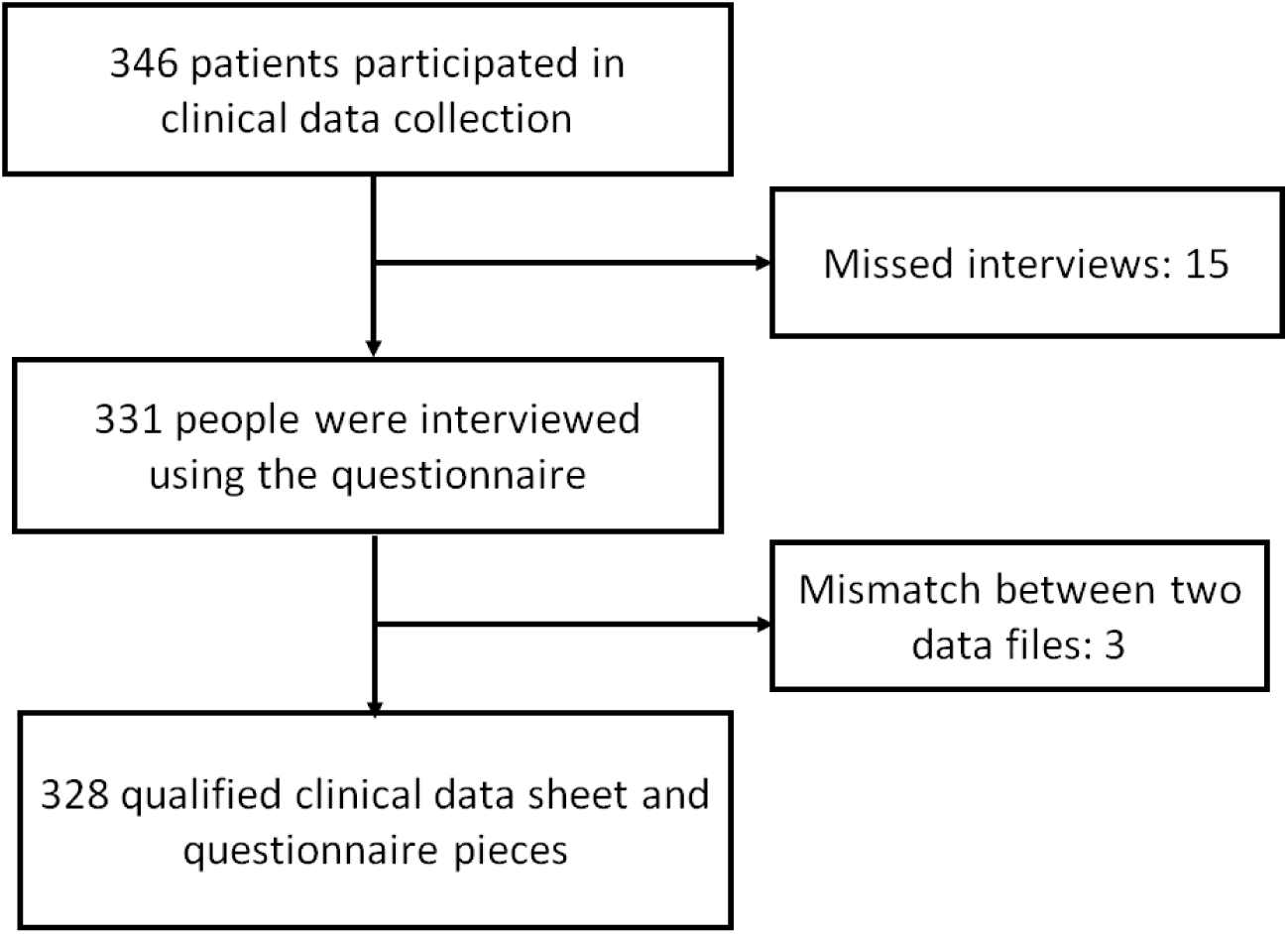
Participants of this study.

### Data collection

The principal investigator and two doctors collected clinical data from medical records using a standardized data form. Assistants (two nursing students and three medical students) were trained for interviews. They collected information from the patients or their relatives on their knowledge and first health-seeking behaviors using a questionnaire.

The clinical data included timeline data (symptom onset, emergency department arrival, neuroimaging, and arrival to the neurology department for definitive treatment), pre-hospital information (location of stroke onset, symptoms, first healthcare seeking, and means of transportation to the first healthcare facilities and Thanh Hoa General Hospital), clinical information at the emergency department (medical history, vital signs, clinical examination results, NIHSS scores, and imaging results), and clinical information at the neurology department (treatment procedures). The study collected data on the starting time of intravenous route placement for thrombolysis (needle time) and groin puncture for endovascular treatment (puncture time) for patients with acute ischemia.

Meanwhile, using a structured questionnaire, the assistants interviewed patients, their relatives, or bystanders (if patients were unconscious). The interviews were completed within one or two working days following admission. The questionnaires collected information on the patient’s age, occupation, education level, measures to support patients at the onset of stroke, and knowledge and awareness about stroke. After completing the questionnaire, the assistants provided interviewees with a leaflet and brief education on stroke, including information on stroke symptoms/signs, appropriate action to support such people, and recurrent stroke prevention.

### Variables

The primary outcome of the study was a pre-hospital delay, which was defined as the duration from stroke symptom onset (or last well-known time for wake-up stroke, as commonly used in previous studies) to the emergency department arrival, defined as the time when the patient underwent a physical examination at the emergency department, as recorded in the medical records. For the analysis, the pre-hospital delay was dichotomized into two categories: ≤ 4.5 hours (early arrival) and > 4.5 hours (delayed arrival). This cutoff was chosen based on the hospital guidelines that recommend providing intravenous thrombolysis within 6 hours of the stroke onset, considering that the in-hospital procedures prior to treatment require 1.5 hours. Secondary outcomes included hospital delay in imaging diagnosis, defined as the duration from arrival at the emergency department to the start of neuroimaging for final diagnosis at Thanh Hoa General Hospital derived from the first image of either CT or MRI, whichever was performed earlier. Another secondary outcome was hospital delay in the definitive treatment, defined as the interval from arrival at the emergency department to transfer to the neurology department for definitive treatment (needle or puncture time was not used because it was not relevant to majority of the patients who were not eligible for thrombolysis or endovascular procedures).

The independent variables included age, gender, education level, occupation, type of public health insurance, affordability to pay for hospital fees, distance from the location where the patient developed the symptom to Thanh Hoa General Hospital (defined as the shortest route calculated using Google Maps and dichotomized into ≤ 10 km and > 10 km), first contact for help, and mode of transportation from home to the first health facility (dichotomized as EMS use and non-use).

To evaluate the knowledge of patients and their family members about stroke, we asked them questions related to four different stroke topics: risk factors (10 questions), symptoms (8), consequences (5), and rehabilitation (4). We created scores for each topic by obtaining the sum of correct answers. The stroke symptom score was used in logistic regression as a 3-category variable: low (1–3), intermediate (4–6), and high (7–8).

### Sample size

The study sample size was calculated using “Sample Size Determination in Health Studies” software version 2.0 (World Health Organization) (19), assuming a comparison of proportions between two independent groups (proportions of delays in non-EMS use and EMS use are approximately assumed to be 0.9 and 0.8 in the two groups, respectively), alpha = 0.05, power = 0.8, the ratio between the two groups = 4 was 375. Considering 20% of the missing data, we needed 450 participants in this study.

### Statistical analysis

Univariate and multivariable logistic regression models were used, with pre-hospital delay as the dependent variable. We selected independent variables based on our hypothesis (EMS use and choice of first medical facilities are important factors), previous findings, and the assumption that demographic factors should be controlled. The selected variables included age, gender, first healthcare-seeking behavior, mode of transportation from the location of stroke onset to the first health facility, distance to Thanh Hoa General Hospital, education level, EMS use, healthcare insurance type, ability to purchase the cost of treatment, knowledge about symptoms at stroke onset, and the type of stroke.

## RESULTS

Among the 328 patients analyzed, 193 (58.8%) were men. The median (IQR) age was 70 years (62–77), and the median distance was 25.1 kilometers (13.65–43). Most patients lived with family members (94.5%), were dependent or retired (60.0%) and had secondary or lower-level education. Most had healthcare insurance (96.9%), but 40 people (12.3%) could not afford the expense (**Table 2**).

**Table 2.** Patients’ sociodemographic characteristics (n = 328)

| **Characteristics** | **Median (IQR) or n (%)** |
| --- | --- |
| Age, y, median (IQR) | 70 (62 – 77) |
| Sex, male, n (%) | 193 (58.8) |
| Distance to hospital, km, median (IQR) | 25.1 (13.65 – 43) |
| Living alone, n (%) | 18 (5.5) |
| Job, n (%) |  |
| Dependent (without pension) | 123 (37.6) |
| Retired | 77 (23.5) |
| Farmer, Forestry and Fishery | 79 (24.2) |
| Freelance workers | 28 (8.6) |
| Service, sales | 8 (2.4) |
| Engineers | 7 (2.1) |
| Other | 6 (1.6) |
| Education level, n (%) |  |
| No education | 14 (4.0) |
| Primary school | 120 (36.7) |
| Secondary school | 116 (35.5) |
| High school | 49 (15.0) |
| College and/or above | 29 (8.9) |
| Public healthcare insurance,* n (%) |  |
| On-line public healthcare insurance | 167 (51.2) |
| Off-line public healthcare insurance | 149 (45.7) |
| Non-public healthcare insurance | 10 (3.1) |
| Unable to purchase treatment cost, n (%) | 40 (12.3) |
| Symptoms when stroke onsets, n (%) |  |
| Hemiparesis, monoparesis, or quadriparesis | 235 (33.5) |
| Aphasia | 154 (21.9) |
| Hemisensory deficits | 103 (14.7) |
| Facial drop | 71 (10.1) |
| Sudden decrease in level of consciousness | 58 (8.3) |
| Vertigo | 38 (5.4) |
| Sudden severe headache | 37 (5.3) |
| Other | 6 (0.8) |
| Patient history, n (%) |  |
| Hypertension | 202 (45.9) |
| There is no recorded clinical history | 76 (17.3) |
| Diabetes mellitus | 58 (13.2) |
| History of stroke | 57 (13.0) |
| Atrial fibrillation | 6 (1.4) |
| Other | 41 (9.4) |
| Diagnosis, n (%) |  |
| Cerebral infarction | 247 (75.3) |
| Cerebral hemorrhage | 81 (24.7) |
| Transportation to Thanh Hoa General Hospital, n (%) | 322 |
| EMS | 8 (2.4) |
| Taxi | 183 (55.8) |
| Private car | 90 (27.4) |
| Motorbike | 34 (10.4) |
| Other | 13 (4.0) |
| Late arrival (Pre-hospital time > 4.5 h), n (%) | 181 (55.4) |
| Prehospital delay, min, median (IQR) | 324 (162.25 – 1091.75) |
| Hospital delay in imaging diagnosis, min, median (IQR) | 14 (10 - 22) |
| Hospital delay in treatment, min, median (IQR) | 66 (40 – 96) |

IQR: interquartile range
\*On-line public healthcare insurance: the healthcare facility that the patients were assigned in their health insurance was Thanh Hoa General Hospital.
Off-line public healthcare insurance: the healthcare facility that the patients were assigned in their health insurance was not Thanh Hoa General Hospital.

Cerebral infarction was the most common type of stroke among the patients, accounting for 75% of the cases. The most frequently reported symptoms during stroke onset were paresis, aphasia, and hemisensory deficit. Hypertension was the most reported medical history, affecting 63.3% of patients, followed by diabetes and stroke history.

One hundred eighty-one patients (55.4%) arrived later than 4.5 h. The median of pre-hospital, imaging, and treatment delays was 324 min (162.25 – 1091.75), 14 min (10–22), and 66 min (40–96), respectively. Twenty-one cases received thrombolysis (6.4%) with a median door-to-needle time of 46 min (IQR, 34 – 98), while four cases received mechanical thrombectomy (1.2%) with a median door-to-puncture time of 100.5 min (IQR, 30.50–412.75)

At symptom onset, most patients arranged their transportation to the hospital without using an ambulance. Some of them provided patients with medicine, food, or drinks (**Table 3**). Only eight people called an ambulance. Other measures included folk remedies, such as massage and finger pricking, calling for help from medical staff near the house, and letting patients rest at home. As the first medical facility, most participants chose public hospitals, Thanh Hoa General Hospital (46.0%), or district hospitals (42.2%). The primary reasons for not calling an ambulance were the lengthy wait for ambulance arrival, unfamiliarity with the phone number for the EMS center, and uncertainty regarding whether ambulance services were available in their area. Overall, respondents had limited knowledge of stroke, including its risk factors, symptoms, consequences, and current rehabilitation therapies.

**Table 3.** Health behaviors after stroke symptom onset among interviewers (patients and their relatives) and knowledge of stroke (n = 328)

| **Health behaviors** | **n (%)** |
| --- | --- |
| Observer's action to support patient,* n (%) |  |
| Calling for transportations to hospital | 287 (55.9) |
| Using medicine | 62 (12.1) |
| Feeding | 10 (1.9) |
| Drinking | 19 (3.7) |
| Calling 115 | 8 (1.6) |
| Other solutions | 119 (23.2) |
| Don't know, don't remember | 8 (1.6) |
| Fist healthcare seeking, n (%) |  |
| Thanh Hoa General hospital | 150 (46.0) |
| District hospital | 136 (42.2) |
| Private hospital | 27 (8.4) |
| Neighboring clinic | 7 (2.2) |
| Pharmacist | 1 (0.3) |
| Traditional healer | 1 (0.3) |
| Reasons for not calling ambulance among non-EMS using participants (n = 320),*† n (%) |  |
| Long waiting time for ambulances | 182 (38.4) |
| Didn't remember/didn't know the ambulance phone number | 96 (20.3) |
| Didn't know that I can call ambulance for those medical emergency events in my location area | 82 (17.3) |
| Hoped that the symptoms will resolve spontaneously | 30 (6.3) |
| Felt unnecessary to call ambulance | 39 (12.3) |
| Feared expenses | 5 (1.1) |
| Other opinions | 40 (8.4) |
| Stroke awareness, points, median (IQR) |  |
| Knowledge about the risk factors of stroke, full score 10 points | 2 (1 – 4) |
| Knowledge about stroke symptoms, full score 8 points | 3 (2 – 4) |
| Knowledge about stroke consequence, full score 5 points | 2 (1 – 4) |
| Knowledge about the rehabilitation therapies, full score 4 points | 1 (0 – 1) |
| Never heard about FAST (signs of stroke onset), n (%) | 298 (91.4) |

IQR, interquartile range; EMS, emergency medical system; FAST, Facial drooping; arm weakness, speech difficulty; time to call ambulance.
\* Multiple choice questions
† This question was answered by 320 respondents who did not use ambulances.

In the univariate logistic regression analysis, long pre-hospital delay was associated with seeking first medical help at district or private hospitals (odds ratio [OR] 1.71, 95% confidence interval [CI] 1.09–2.68), location distance farther than 10 km (OR 2.02, 95% CI 1.15–3.56), and lower education level (Table 4). EMS use, stroke symptom knowledge, living alone, and stroke type were not associated with pre-hospital delay. The multivariable analysis showed that longer pre-hospital delay was significantly associated with location distance > 10 km (OR 2.07, 95% CI 1.09–3.95) and secondary education level (OR 1.99, 95% CI 1.06–3.73) after adjusting for the other factors. Seeking first medical help at district and private hospitals did not show a significant association, although the OR was 1.57 (95% CI, 0.93–2.65).

**Table 4.** Multivariate logistic regression model investigating the association between patient characteristics and late arrival (> 4.5 h) after stroke onset (n = 328)

|  | Crude odds ratio (95% confidence interval) | Adjusted odds ratio (95% confidence interval) |
| --- | --- | --- |
| Age, years |  |  |
| ≤ 44 | 1 | 1 |
| 45 – 54 | 0.58 (0.13 – 2.51) | 0.77 (0.16 – 3.70) |
| 55 – 64 | 0.73 (0.19 – 2.79) | 1.22 (0.28 – 5.39) |
| 65 – 74 | 0.72 (0.19 – 2.68) | 1.38 (0.32 – 5.99) |
| ≥ 75 | 1.15 (0.31 – 4.32) | 1.87 (0.43 – 8.17) |
| Gender (Male/Female) | 1.16 (0.75 – 1.81) | 1.17 (0.71 – 1.93) |
| Distance |  |  |
| ≤ 10 km | 1 | 1 |
| > 10 km | 2.02 (1.15 – 3.56) | 2.07 (1.09 – 3.95) |
| Time (Night/Day) | 0.60 (0.37 – 0.98) | 1.63 (0.97 – 2.75) |
| Living alone (Yes/No) | 2.02 (0.76 – 5.36) | 2.33 (0.77 – 7.05) |
| Education level |  |  |
| No education/Primary education | 1.83 (1.04 – 3.23) | 1.82 (0.98 – 3.37) |
| Secondary education | 1.90 (1.06 – 3.40) | 1.99 (1.06 – 3.73) |
| High school education and/ above | 1 | 1 |
| First healthcare seeking |  |  |
| Neighboring clinic, pharmacists,<br>traditional healers | 0.52 (0.12 – 2.16) | 0.51 (0.11 – 2.28) |
| District hospital and private<br>hospital | 1.71 (1.09 – 2.68) | 1.57 (0.93 – 2.65) |
| Thanh Hoa General Hospital | 1 | 1 |
| EMS using (No/Yes) | 2.52 (0.62 – 10.25) | 2.11 (0.46 – 9.71) |
| Public healthcare insurance |  |  |
| On-line insurance | 1 | 1 |
| Out-line insurance | 0.95 (0.61 – 1.48) | 0.94 (0.58 – 1.53) |
| Non healthcare insurance | 0.78 (0.22 – 2.78) | 1.00 (0.25 – 4.10) |
| Ability of purchase the cost of<br>treatment (No/Yes) | 1.57 (0.79 – 3.13) | 1.71 (0.79 – 3.70) |
| Knowledge about symptoms when<br>stroke onsets |  |  |
| Low (1 – 3) | 1.04 (0.49 – 2.19) | 1.09 (0.47 – 2.57) |
| Mediate (4 – 6) | 1.34 (0.58 – 3.07) | 1.37 (0.53 – 3.40) |
| High (7 – 8) | 1 | 1 |
| Type of stroke (infarction /<br>hemorrhage) | 0.83 (0.49 – 1.36) | 0.92 (0.52 – 1.65) |

EMS, emergency medical system

## DISCUSSION

This study indicated that most patients with stroke in the Thanh Hoa Province in Vietnam did not reach the hospital on time. Given the relatively short hospital delay, the pre-hospital delay mostly contributed to the total time loss. First medical seeking did not show any significant association by a very small margin in the multivariate analysis. The other hypothesized factor, ambulance use, was not found to be significantly associated. The factors associated with pre-hospital delay included long distance from the hospital and low education level.

Studies have consistently shown that long travel distance is associated with high odds of delayed hospital arrival. (20, 21) Moreover, poor road conditions and limited availability of transportation exacerbate the challenging accessibility to hospitals in remote areas. Given that the Thanh Hoa General Hospital is the sole stroke center in the province, patients residing in remote regions may have to travel extended distances to reach there.

Patients’ first choice of seeking medical help may be an important factor. If they opt for district or private hospitals that cannot diagnose or treat stroke, they may have to be transferred to the stroke center without receiving any treatment, leading to a waste of time.(18, 21) Despite the significance in the univariate analysis, the multivariable analysis did not demonstrate a significant association between pre-hospital delay and first medical help-seeking by a small margin. This may be due to the limited sample size, and a larger sample size may have yielded statistical significance. Conversely, seeking medical help at clinics, pharmacists, or traditional healers was not associated with delay, possibly because they advised prompt seeking medial help at the tertiary care hospital.

Although we hypothesized that patients who used ambulances would arrive at the hospital earlier, the logistic regression model did not show any association between EMS use and pre-hospital delay; although the odds of delay were two times higher among EMS non-users than users. The small number of EMS users in the study (only eight) may have limited the statistical power, leading to the lack of significance. Therefore, further research with a larger sample size is needed to answer the question of whether EMS use can reduce pre-hospital delay in immature EMS systems.

Although insignificant in this study, EMS remains important in promptly transporting patients with stroke. Of the eight EMS users in the study, all were directly transported to the provincial hospital. Because choosing district or private hospitals as the first choice for medical help may increase the prehospital delay, the EMS crews’ decisions were appropriate.

This study also highlights the limited knowledge of stroke among patients and their family members. Previous studies have shown a significant association between knowledge about stroke and pre-hospital delays. (18) However, in this study, there was no association between knowledge of symptoms at stroke onset and pre-hospital delay. This may be because the study population had uniformly low levels of knowledge about stroke symptoms (median score was very low), which may have limited the ability to detect significant associations with the outcome.

Reasons for not using the ambulance included long waiting times, and lack of knowledge of the EMS system, including the when and how to use it. Combined with the finding of insufficient knowledge of stroke, the short-term possible intervention should be awareness-raising campaigns. Such campaigns should be focusing on “be aware of stroke symptoms” and “directly go to the stroke center” using the fastest transportation available.

Long-term interventions should include expansion of EMS coverage and upgrading the capacities of district hospitals to diagnose and treat stroke. We may adopt either “Drip-and-Ship” or “Mothership” strategies according to the distance to Thanh Hoa General Hospital. (22, 23) In remote areas where the travel time is greater than 30 minutes, thrombolysis should start at the nearest district hospital, and the patients should immediately be transferred to Thanh Hoa General Hospital (drip-and-ship). In areas within a 30-min drive, patients should be directly transported to Thanh Hoa General Hospital (mothership). (24)

In-hospital delay in imaging and treatment in this study was not very long compared to the recommended time: 10 min or less for imaging and 30 min for thrombolytic treatment (to achieve a system where 95% of patients are treated within 60 min, the median door-to-needle time should be 30 min). (25) The current system in which patients with stroke receive treatment at the neurology department can be modified to receive thrombolytic treatment even at the emergency department while waiting for their transfer to the neurology department. The use of CT/MRI or angiography rooms should be more flexible so that emergency cases can be promptly accepted. (26)

This study provides novel insights into stroke management in Thanh Hoa Province and Vietnam’s emergency healthcare system. As the first study to investigate the emergency system’s role in transporting patients with stroke, it sheds light on the region’s stroke treatment landscape. The prospective design enhances data reliability and reduces information omission. These findings can potentially improve both pre-hospital and in-hospital stroke management in Thanh Hoa Province and other stroke centers.

However, several limitations to this study should be acknowledged. First, the sample size was not large enough, which resulted in some findings falling short of statistical significance by a small margin. A more extensive study is required to confirm the role of EMS in facilitating access to medical care for patients with stroke. Second, due to the limited workforce for data collection, some eligible patients may have been missed during the study period, particularly at night, which could have introduced selection bias. However, we believe that missing patients at night would not have significantly impacted our findings since pre-hospital and in-hospital delays are often worse during these times. Third, this study did not identify specific in-hospital factors contributing to delays in imaging and treatment, which should be analyzed in further studies.

## Conclusions

The study revealed that many patients with stroke failed to reach the hospital in a timely manner to receive recanalization therapy and awareness of stroke among themselves and their relatives were poor. Patients’ first healthcare seeking whether to go directly to the provincial hospital or EMS use was not significantly associated with pre-hospital delay. Further research is needed to investigate the reasons for pre-hospital delays and develop targeted interventions to improve stroke awareness and reduce the delays.

## Data Availability

Data are available from the corresponding author upon reasonable request.

